# Community-based models of care for management of type 2 diabetes mellitus among non-pregnant adults in sub-Saharan Africa: a scoping review

**DOI:** 10.1101/2022.11.17.22282376

**Authors:** Emmanuel Firima, Lucia Gonzalez, Fabiola Ursprung, Elena Robinson, Jacqueline Huber, Jennifer M. Belus, Fabian Raeber, Ravi Gupta, Gibrilla F. Deen, Alain Amstutz, Bailah Leigh, Maja Weisser, Niklaus Daniel Labhardt

**Author notes:** Corresponding author: Niklaus D. Labhardt, Div. Clinical Epidemiology, University Hospital Basel, Totengaesslein 3, 4051 Basel, Switzerland. Alternative Corresponding author: Emmanuel Firima, Swiss Tropical and Public Health Institute, Kreuzstrasse 2, 4234 Allschwil.

## Abstract

**Introduction:** The prevalence of type 2 diabetes mellitus (T2DM) and associated morbidity and mortality are increasing in sub-Saharan Africa (SSA). To facilitate access to quality care and improve treatment outcomes, there is a need for innovative community care models and optimized use of non-physician healthcare workers bringing diagnosis and care closer to patients’ homes.

**Aim:** We aimed to describe with a scoping review different models of community-based care for non-pregnant adults with T2DM in SSA, and to synthesize the model outcomes in terms of engagement in care, blood sugar control, acceptability, and end-organ damage. We further aimed to critically appraise the different models of care and compare community-based to facility-based care if data were available.

**Methods:** We searched Medline, Embase, Cumulative Index to Nursing and Allied Health Literature (CINAHL) and Scopus, supplemented with backward and forward citation searches. We included cohort studies, randomized trials and case-control studies that reported on non-pregnant individuals diagnosed with T2DM in SSA, who received a substantial part of care in the community. Only studies which reported at least one of our outcomes of interest were included. A narrative analysis was conducted, and comparisons made between community-based and facility-based models, where within-study comparison was reported.

**Results:** 5,335 unique studies were retrieved, four of which met our inclusion criteria. Most studies were excluded because interventions were facility-based; community care interventions described in the studies were add-on features of a primarily facility-based care; and studies did not report outcomes of interest. The included studies reported on a total of 383 individuals with T2DM. Three different community care models were identified. 1) A community-initiated model where diagnosis, treatment and monitoring occurred primarily in the community. This model reported a higher linkage and engagement in care at 9 months compared to the corresponding facility model, but only slight reductions of average blood glucose levels at six months compared to baseline. 2) A facility-originated community model where after treatment initiation, a substantial part of follow-up was offered at community level. Two studies reported such a model of care, both had as core component home-delivery of medication. Acceptability of this approach was high. But neither study found improved T2DM control when compared to facility care 3) An eHealth model with high acceptability scores for both patients and care providers, and an absolute 1.76% reduction in average HbA1c levels at two months compared to baseline. There were no reported outcomes on end-organ damage. All four studies were rated as being at high risk for bias.

**Conclusion:** Evidence on models of care for persons with T2DM in SSA where a substantial part of care is shifted to the community is scant. Whereas available literature indicates high acceptability of community-based care, we found no conclusive data on their effectiveness in controlling blood sugar and preventing complications. Evidence from larger scale studies, ideally randomized trials with clinically relevant endpoints is needed before roll-out of community-based T2DM care can be recommended in SSA.

## Introduction

Diabetes mellitus (DM) is one of the most common chronic diseases worldwide^1,2,3,4^, and majority of patients present with type 2 DM (T2DM)^5,6^. In sub-Saharan Africa (SSA), about 24 million people currently live with DM and this number is expected to more than double to 55 million by 2045^4,7^. End-organ complications like retinopathy, peripheral sensory neuropathy, nephropathy, and cerebrovascular accidents contribute to mortality and disability^5,8-10^. Traditionally, management of patients with diabetes in sub-Saharan Africa is carried out in health facilities^11^, or with occasional community linkages as ‘add-on’ service^12^. Patients within this care model go through clinics that are often congested, distant from their homes or working places, and have to wait long hours to access care^11^. High direct and indirect costs are majorly borne by patients as out-of-pocket payments^13^. Poor access to care as a result of traditional models of care and rising costs have led to under-diagnosis, under-treatment and consequently poor health outcomes for people living with T2DM in these settings.^11,13^

The Sustainable Development Goals set to reduce by 2030 the burden of non-communicable diseases, and achieve universal health coverage^14^. To meet these ambitious targets, health models that increase access to care especially in low-resource regions like SSA must be developed, validated, and scaled-up^15^. Such models will need to reduce cost, be acceptable as well as feasible. Although primary care centers (PHCs) could potentially fill this gap, decentralizing DM care to PHCs has been sub-optimal, with unsatisfactory outcomes^16,17^. As such, health models that merely introduce community linkages as ‘add-ons’ to primarily facility care models will not be enough, rather community models that effectively reduce the frequency of patient contact with health facilities to reduce cost to patients, and cost and workload at the clinics.^15,18,19^

Community-based care refers to interventions delivered outside of formal health facilities^12,19^. It includes the services of professionals in residential and community settings in support of self-care, home-care, long-term-care, and treatment for substance use disorders and other types of health and social care service^19^. A systematic review assessing studies in different low and middle-income settings showed the usefulness of community-based programs to improve outcomes in immunization programmes, uptake of breast feeding and adherence to tuberculosis treatment^20^. Another systematic review and meta-analysis on the effect of community-based programs on diabetes prevention in low- and middle-income countries^1^ revealed that such programs had positive outcomes on patients at risk of T2DM. A recent scoping review indicates substantial potential of community-based care models for arterial hypertension in SSA ^21^. Moving care of uncomplicated cases and low-risk groups to the community level and to non-physician health workers who are themselves supervised by higher health cadres, has advantages including fewer clinic visits, not having to travel long distances, not waiting in queues, and freeing up medical services in the facility for complicated cases and high-risk groups. This has the potential to provide effective care to both the groups receiving care in the community and those receiving care in the facility^22^.

Currently, there is a lack of evidence on the effectiveness of community-based care for management of DM in the SSA region. We conducted a scoping review to map currently existing models of T2DM community-based care among non-pregnant adults in SSA; synthesize evidence on clinical outcomes of those care models in terms of engagement in care, blood sugar control, end-organ damage, as well as acceptability to both patients and care providers; and to compare the performance of the community-based models of care to facility-based care, if reported.

## Materials and Methods

We conducted this scoping review using the framework initially developed by Arskey and O’Malley, and further refined by Levac et al. and the Joanna Briggs Institute^23-25^. The study protocol with detailed description of our method has been published^26^. Briefly, we searched Medline, Embase, Cumulative Index to Nursing and Allied Health Literature (CINAHL) and Scopus on 23^rd^ May 2021 and 15^th^ October 2021 using the following keywords: “community-based care”, “type 2 diabetes” and “sub-Saharan Africa”. Our final search was conducted on the 24^th^ October 2022 to update the first search. (The search string is available on Supplementary file, S1 table). If screened articles described study protocols that were topically relevant, first authors of those articles were contacted for any initial data on their studies. Forward and backward citation searches were carried out on articles that were included after full text screening.

We included studies carried out in SSA which reported community models of care where the majority of care was delivered outside of, and reduced frequency of patient contact with, traditional health facilities. In studies where care delivered in the community did not reduce contact with health facilities, such community-based care was considered ‘add on’ care, and the studies excluded. We only included studies that reported at least one of the following outcomes: engagement in care, blood glucose indices, T2DM complications, or acceptability of care to patients and providers. See table 1 for PICO framework. Since definition of engagement in care differed between studies, we adopted the definition used in the respective study. T2DM complications included development of retinopathy, neuropathy, nephropathy or diabetic foot syndrome. Acceptability of care was the uptake and utilization of the models of care by the patients or healthcare providers. As acceptability of care is variously defined^27^, we adopted scales used by authors of the respective study. There was no restriction on language. We excluded studies on pregnant women and patients below 18 years of age. We included studies that were prospective or retrospective cohorts, randomised controlled trials, non-randomised controlled trials, and quasi-randomised controlled trials. See Supplementary file, S2 table and S3 table.

**Table 1.** PICO framework

| **Criteria** | **Determinants** |
| --- | --- |
| Population | Adult persons with non-gestational type 2 diabetes mellitus in sub-Saharan Africa |
| Intervention | Community-based care delivery |
| Comparison | Facility-based care (where available) |
| Outcome | Acceptability to patients and/or healthcare workers, Fasting blood glucose, Random blood glucose, glycated haemoglobin (HbA1c), engagement in care, development of T2DM-related complications |

All search results identified using respective search strings for Medline, Embase, Cumulative Index to Nursing and Allied Health Literature (CINAHL) and Scopus were imported into EndNote™ and de-duplicated. Initially, two reviewers (EF and FU) independently screened all abstracts, applying the pre-defined eligibility criteria. Abstracts were excluded if they did not meet our inclusion criteria; or included for full text screening if they either met our inclusion criteria or if eligibility could not be determined immediately. Afterwards, full texts of all included studies were retrieved. Reviewers (EF and FU) independently screened the full texts for inclusion. Any disagreements were resolved by discussions between EF, LGF and NDL. Studies which were initially included but excluded during screening of the full text were specifically labelled as such in a table of excluded studies including the reason for exclusion.

A data extraction tool was created in Word™ and designed to collect information on author, year of publication, study design, location of study, duration of follow-up, type of community-based care model, health provider cadre, special trainings administered to providers and outcomes assessed. Where applicable, outcomes in a comparator arm (facility-based care) were also extracted (see Supplementary file, S4 table). Data extraction was done independently and in duplicate by EF and FU. Discrepancies were discussed and resolved in consultation with a third person (NDL).

A textual narrative synthesis approach was used for analysis and synthesis^28^. In a first step, we identified and classified the model(s) evaluated in each study. For this, we used the framework on primary care-based models of NCD care in SSA by Kane et. al^29^. This framework classifies models of care according to origin or source of included patients; key activities undertaken within the care model; key cadre of participating staff; additional staff preparation for model; integration with other care; follow up and evaluation plan; and outcome. Afterwards, the components of each model were summarized. Findings are presented using tables and narrative reporting.

Quality of the cohort studies was assessed using the Newscatle-Ottawa scale^30,31^. The scale grades selection, comparability and outcome domains to an overall maximum score of 9. Although thresholds are not validated, we adopted the approach proposed in a recent review where scores of less than 6 are considered to be of high risk of bias.^32^ We assessed the randomized controlled trial using the Cochrane Collaboration’s tool for assessing risk of bias in randomized controlled trials.^33^ We assessed bias in the randomization process, deviation from intended intervention, completeness of outcome data for each main outcome, bias in the measurement of outcome and bias in the selection of the reported result.^34^

## Results

After de-duplication, our database search yielded 5,335 records. Additionally, we retrieved 164 articles from backward and forward citation search. We assessed 83 full text articles out of which 4 articles met our inclusion criteria and are included in this scoping review. See PRISMA^35^ diagram in figure 1. Reasons for exclusion were: non-eligible patient population (pre-diabetes, pregnant women, type 1 diabetes); primarily facility-based model for delivering treatment with community-based model as add-on; incomplete description about the main characteristics needed to define the model; not reporting relevant outcomes; non-eligible study design. Numbers of studies excluded for each reason are presented on the flow chart in figure 1.

**Figure 1.**
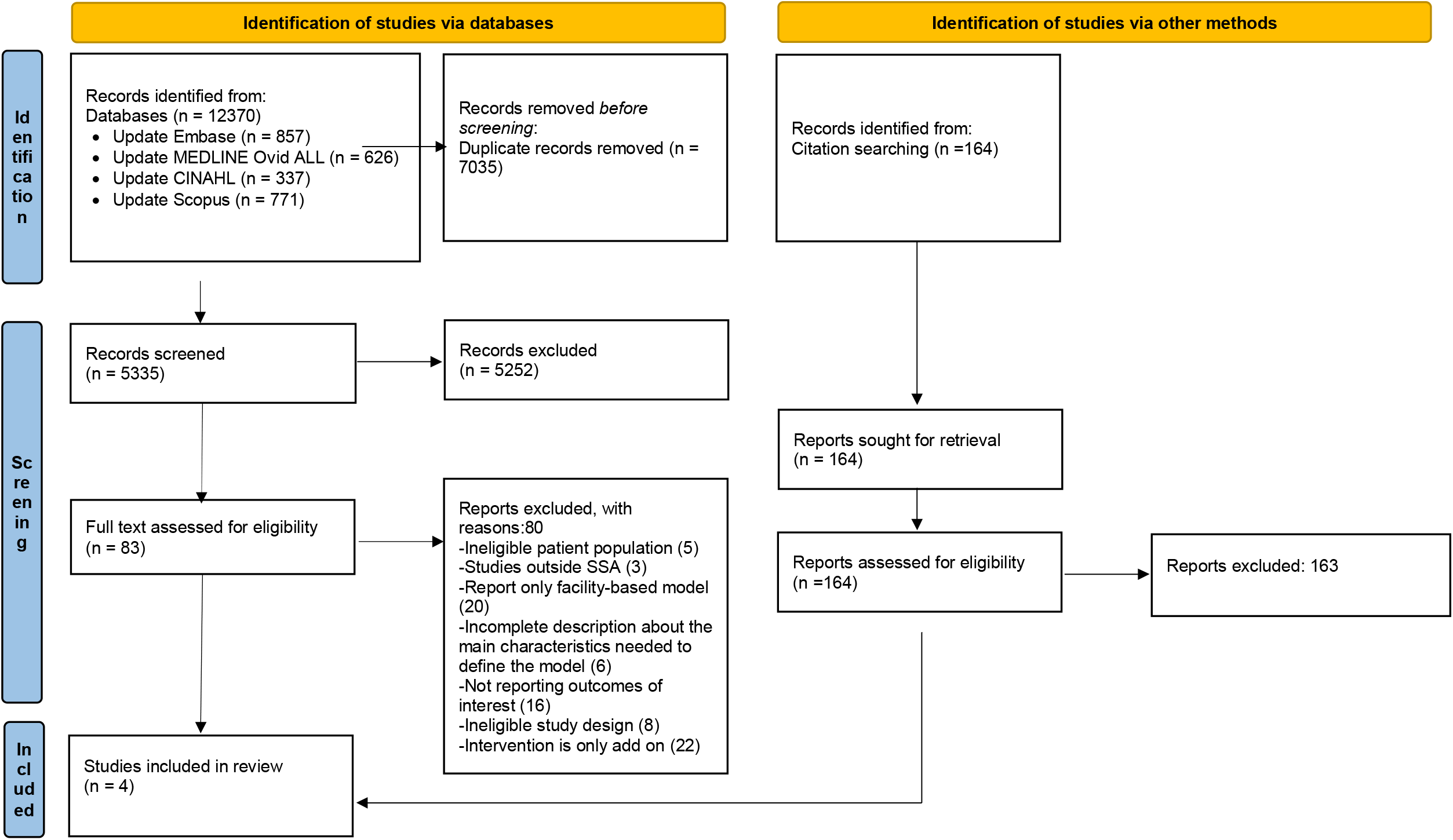
Flow chart showing results of database search and screening of primary articles.

### Study characteristics

Characteristics of the four studies including design and intervention are summarized on table 2. The first study was a mixed-method study conducted in an urban setting in South Africa^36^. The quantitative aspect of this study was a prospective cohort with a non-randomly selected control group. The second study was an observational cohort study with a historical control group conducted in a rural setting in Kenya^37^. The third study was a randomized pilot trial without formal comparison between the two groups conducted in an urban area in the Democratic Republic of Congo^38^. The fourth study was a retrospective cohort study with a matched control group, conducted in an urban area in South Africa^20^. Overall, the four studies report on *N*=383 T2DM patients within the community care models.

**Table 2.**
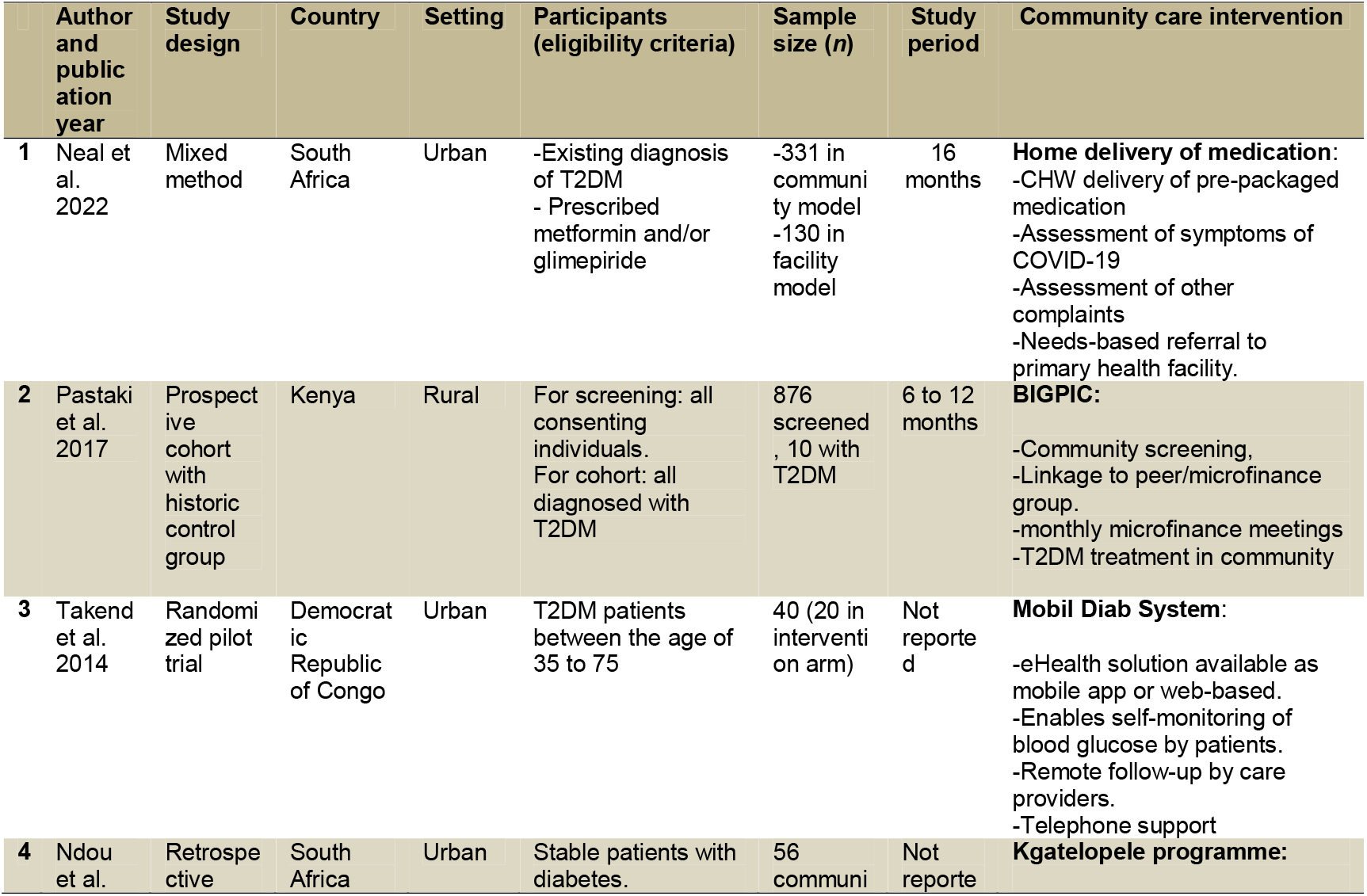
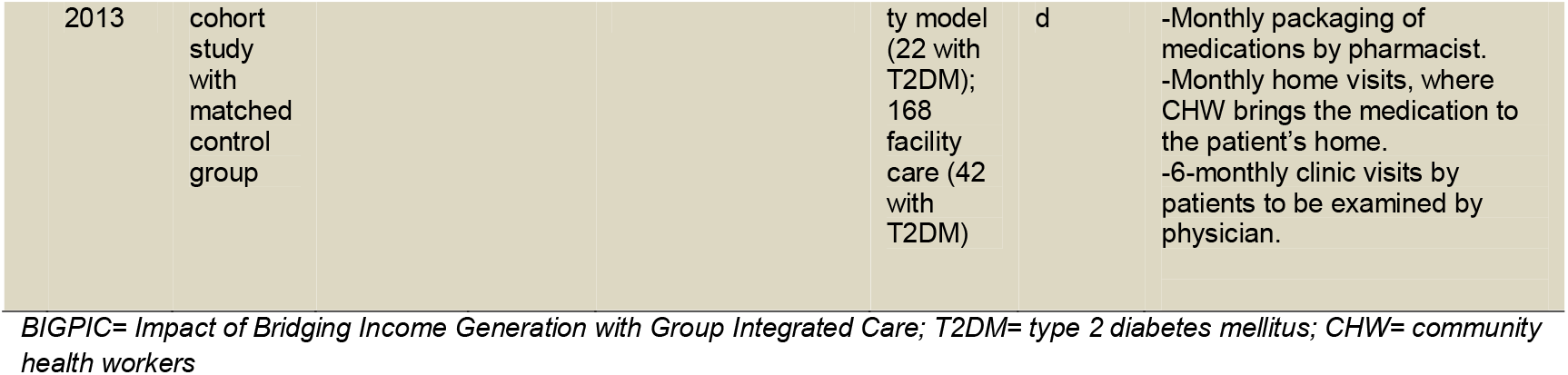
Summary of main study characteristics

### Models of Care

Based on the framework by Kane et. al^29^, we identified three models of care: community-initiated model; eHealth model; and facility-originated community model (table 3).

**Table 3.** Main components of the community care interventions described presented by care model categories.

| Community care intervention | Source of patients | Location of care | Key activities undertaken | Cadre of staff used | Additional staff preparation for model delivery | Integrated care | Outcome measured |
| --- | --- | --- | --- | --- | --- | --- | --- |
| **Community-initiated model** |  |  |  |  |  |  |  |
| BIGPIC | Community-based screening, enrolment of individuals diagnosed with diabetes | Households and places used for gatherings like religious centers, schools, markets and shops | Medical prescription and drug distribution, lifestyle change support | Community care workers<sup>a</sup>, local volunteers, social workers, pharmacists, clinical officers, and physicians | Not reported | Health education with agro-business advice<br>economic stability through loans within microfinance groups<br>Demand for care creation through incentives<br>-aHT co-management | Blood glucose, HbA1c<br>Linkage to care<br>Engagement in care |
| **eHealth model** |  |  |  |  |  |  |  |
| Mobil Diab System | Known T2DM patients, source of recruitment not reported | Long distance support using eHealth tools e. g specially deployed app | Blood glucose monitoring by patient and reported through eHealth tool | Physician<sup>b</sup> | Not reported | None | HbA1c,<br>Acceptability to patient<br>Acceptability to healthcare provider |
| **Facility-originated community model** |  |  |  |  |  |  |  |
| Home delivery of medication | Primary health centers | Patient's household | Medication prescription and distribution | community health worker<sup>a</sup> |  | Co-management with aHT | HbA1c,<br>Acceptability to patient |
| Kgatelopele programme | Primary health centers | Patient's household | Medication prescription and distribution to patient's home. Health promotion, education, adherence counselling. | Physician, pharmacist, community health worker<sup>a</sup> | 14-week training for CHW on home care, adherence counselling and health promotion | Co-management with aHT | Blood glucose |

*BIGPIC= Impact of Bridging Income Generation with Group Integrated Care; T2DM= type 2 diabetes mellitus; aHT= arterial hypertension*
<sup>a</sup>Main staff cadre providing services.
<sup>b</sup>Self-care driven, with physician supervision

Within the community-initiated model of care, patients were actively sought in the community via various methods of screening and diagnosis, followed by within-community treatment initiation and monitoring. Using this model was the Impact of Bridging Income Generation with Group Integrated Care (BIGPIC)^37^ intervention which was developed in rural Kenya and focused on diabetes and hypertension care. The intervention included community-based screening and diagnosis; linkage to a peer group and a microfinance group; integration of health education and business counselling; incentives to generate demand for care; and care provision in the community. The study reported on blood glucose, linkage to and engagement in care. It compared linkage to, and engagement in care with a historical facility-based comparison group. In BIGPIC, primarily non-physician clinicians screened, diagnosed and treated patients in the community either at or near the patient’s home. An important additional component of BIGPIC was integration of diabetes care with hypertension care and economic empowerment.

The Mobile Diab System^38^, an e-health model used for patients already diagnosed and treated for T2DM, was implemented in two urban areas of the Democratic Republic of Congo. This model aimed to improve access to care, reduce frequency of clinic visits and increase patient’s involvement. With this model, patients were able to self-monitor their health, for example track exercises, drug intake and blood glucose measurement. These data were then sent via a portal to the patient’s physician who reviewed the information. Feedback, including therapy adjustments, instructions or recommendations were sent back to patients through the mobile system. Thus, central to this model were long-term medication prescription and distribution, self-care, self-monitoring and clinician follow-up via eHealth platforms. The study reported on blood glucose levels, HbA1c as well as acceptability of the care model to both patients and care providers. The model empowered patients, ensured information flow between providers and patients, and reduced the frequency of clinic visits.

In the facility-originated model, patients who received care at a facility were transferred to community-based follow-up. Two studies fell into this model, the Kgatelopele programme and the Home Delivery of Medication (HDM) intervention. The Kgatelopele programme^20^ sought to improve care for people living with diabetes and hypertension by providing home care through community health workers. The overall aim was to improve acceptability, accessibility, and affordability of care.

Activities undertaken were monthly packaging of medications by pharmacists, monthly home visits by community health workers who then brought the packed medications to patients, and six-monthly clinic visits. Although the main staff cadre involved was the community health worker, this programme included specialized care from pharmacists and physicians at the clinic. Community health workers also provided social support geared towards improved patient literacy about their condition, adherence to medication and clinic visits. Patients were recruited at the clinic and outcomes were retrospectively compared to a matched cohort from the same clinic who did not enroll in the community programme. The Kgatelopele programme was similar to the community-initiated model BIGPIC, with the main exception that it was a facility-originated community model where known and stable patients were transferred from clinic-based care to community care. As in the community-initiated model, care and medication were delivered by non-physician health workers. Similar to BIGPIC, the Kgatelopele programme integrated T2DM management with care for hypertension.

Home Delivery of Medication(HDM)^36^ intervention was developed due to difficulties faced in health facilities during the Coronavirus disease (COVID-19) pandemic. The aim of the intervention was to determine if patient follow-up in the community with HDM improved blood glucose control as shown in HbA1c levels, and acceptability by patients. Community health workers delivered pre-packaged medications to eligible patients. They also performed various evaluations including assessment for COVID-19 symptoms and other complaints with referral to primary health facilities if needed.

### Reported outcomes and comparisons to facility-based model of care

The BIGPIC study from Kenya reported on linkage to care, engagement in care, blood glucose and HbA1c. The Mobil Diab pilot trial from the Democratic Republic of Congo reported on acceptability by patients and providers, blood glucose values and HbA1c, and the Kgatelopele study reported blood glucose values only. The HDM intervention reported on HbA1c and acceptability by patients. There were no reported outcomes on end-organ damage. See table 4.

**Table 4.** Summary of outcomes and comparisons where relevant

| Author and publication year | Study period/timing of outcome | Model | Acceptability |  | Blood sugar level* |  | Linkage and engagement in care |  |
| --- | --- | --- | --- | --- | --- | --- | --- | --- |
|  |  |  | Model outcome | Conventional care outcome | Model outcome | Conventional care outcome | Model outcome | Conventional care outcome |
| **Pastakia et al. 2017** | 6 to 12 months | Community-initiated | - | - | FBS: 160.4mg/dl at baseline; 153.2mg/dl after 6 months.<br>HbA1c: 10.8% at baseline; 10.0% after 6 months | - | 100% 10/10 with T2DM linked to care<br>70% engagement in care at 9 months | 31% linkage to care |
| **Takeda et al. 2014** | 2 months | eHealth | For patients: Mean score of 8.65 on a scale of 1 to 10.<br>For medical staff: Mean score of 8.75 | - | HbA1c: 8.65% at baseline; 6.89% at 2 months | HbA1c: 8.59% at baseline; 8.6% at 2 months | - | - |
| **Ndou et al. 2013** | NR | Facility-originated community model | - | - | FBS 3.6-5.8 mmol/L: 9.1% | FBS 3.6-5.8mmol/L: 26.1% | - | - |
| **Neal et al. 2022** | 16 months | Facility-originated community model | 100% acceptability of model | - | HbA1c: 9.3% at baseline; 9.5% at follow-up<br>28% of patients with controlled<sup>#</sup> DM at baseline; 27% of patients with controlled DM at follow-up | HbA1c: 9.6% at baseline; 10.1% at follow-up<br>27% of patients with controlled DM at baseline; 22% of patients with controlled DM at follow-up |  |  |

NR= not reported; FBS= fasting blood sugar; HbA1c= glycated haemoglobin. \* Reported FBS and HbA1c values are averages.
<sup>#</sup> Controlled DM for the intervention was HbA1c < 7.5%

#### Engagement in care

Linkage to care, which was defined as return to subsequent group meeting following positive screening for T2DM, was 100% for BIGPIC’s community model compared to 31% in the historic conventional care group. At 9 months seven (70%) of the 10 T2DM patients were still in care. Linkage to care, and engagement in care were reported only in BIGPIC’s community care model.

#### Blood glucose control

In the Kgatelopele program, blood glucose control was defined as a fasting blood glucose of between 3.6 to 5.8 mmol/L among people with T2DM. Applying this definition, 2/22 (9%) and 11/42 (26%) achieved control in the community and the facility care group, respectively. In BIGPIC’s community-initiated model, average fasting blood glucose in the 10 patients with T2DM was 160.4mg/dL (8.9 mmol/L) at baseline and reduced to 153.2mg/dL (8.5 mmol/L) after 6 months representing a non-significant reduction. Average HbA1c decreased from 10.8% at baseline to 10.0% after 6 months. Blood glucose indices were not reported for the historical control group. Within the Mobil Diab system’s eHealth model, baseline average HbA1c was 8.65% with a 1.76 percentage points reduction to 6.89% at 2 months. This model was compared to conventional care where at 2 months average HbA1c remained unchanged (baseline 8.59%, 2-months 8.6%). For the HDM intervention, average baseline HbA1c values were high for both HDM cohort and non-HDM cohort. These values increased at follow-up, but with a non-significantly lower increase within HDM cohort compared to non-HDM cohort. There was a 1% decrease in controlled DM at follow-up within HDM cohort whereas number of patients with controlled DM reduced by 5% within non-HDM cohort.

#### Acceptability

Acceptability was reported by both HDM intervention and Mobil Diab system’s eHealth model. For the HDM intervention, implemented during the COVID-19 pandemic, patients’ acceptability was assessed by questionnaires. All interviewed patients preferred HDM because it was convenient, safe, resolved their transport difficulties, and reduced their fear of getting infected in the facility. Within Mobil Diab system’s eHealth model, acceptability to patients and acceptability to healthcare providers were described on a 10-point scale based on responses to two questions, each directed to patients and to healthcare providers. The questions explored whether patients and healthcare providers would wish to continue using the system, and whether they would recommend the system to others. The average score to both questions was then taken as the acceptability score. The scores were 8.65 and 8.75 out of 10 for patients and healthcare workers respectively. These results, according to the authors, reflected an overall positive acceptance of the system by the patients and health workers. The authors also stated that one reason for patient acceptance of the model was the motivation to reach target blood glucose levels since patients could easily access the database which showed carbohydrate contents of the meals they were taking. For medical practitioners, the system was suitable because of the possibility to supervise more patients simultaneously and remotely. The major concern for both patients and health workers were internet cost and time required to get familiar with both mobile and web applications. Suggestions to improve acceptability and performance of the eHealth platform included updating the drug and food list in the apps to include more locally available items, making available more trainings about use of the system, introducing glucose measuring device kit, and creating within the model an avenue for sporting activities for patients.

### Quality of evidence

HDM intervention, BIGPIC and Kgatelopele cohort studies scored below 6, thus classified as being of high risk of bias. The low scores were mainly due to a lack of comparators (BIGPIC) or non-randomly selected comparators (Kgatelopele programme). See Supplementary file, S5 table. The included randomized pilot trial had overall high risk of bias. The randomization domain was found to be of high risk of bias. There were some concerns related to deviation from intended intervention, measurement of the outcome and selection of the reported result. See Supplementary file, S6 table. Further, reporting of the trial did not follow standards for randomized controlled trials^39^.

## Discussion

We conducted a scoping review to describe published data from cohort and intervention studies that assessed community-based models of care for people living with T2DM in SSA where a relevant part of care was provided outside the facility. To our knowledge, this is the first review that addresses this question. Our literature search strategy yielded four eligible articles out of 5,335 distinct records. The four studies were very heterogeneous in design, type of care model described, and outcomes reported. Quality of evidence provided by the studies as well as the relatively low number of T2DM patients enrolled in the four community models of care make it impossible to conclude on the effectiveness of community-based T2DM care in SSA. As such, this scoping review’s main finding is that there is a considerable evidence gap regarding community-based T2DM care in SSA, and larger scale, well designed studies are needed. Such future studies may build on the experience reported in the four studies included in this review.

The Kgatelopele’s programme, a facility initiated community model of care, reported only on blood glucose levels. After treatment initiation at the facility, significant aspects of patient care were moved to the community for 22 selected patients among whom only two achieved blood sugar control. In contrast, 46 of the 168 (26%) remaining in facility-based care achieved glycemic control. In BIGPIC’s community-initiated model, with 10 T2DM patients, there was only a very modest reduction in fasting blood glucose and HbA1c at six months follow-up. Similarly, the HDM Intervention found no clear indication of better T2DM control in the cohort receiving home delivered medication. These results differ from other reports from LMICs^1^ which showed significant reductions in fasting blood glucose and HbA1c measurements in favor of community-based models care where the community-aspect was an add-on to and did not replace standard facility care.

In one of the studies assessed in our full text review but excluded from the scoping review^40^, the intervention supported self-monitoring of blood glucose and relied on six approaches: identification of high risk patients by clinicians and enrollment in the intervention by community health workers; sending patients home with a glucometer and cell phone access; weekly follow-up via phone calls for blood glucose results; glucose results and medication dose summaries generated for clinician to review. Although utilizing an eHealth model for follow-up, the study was not included in this review as majority of participants had type 1 diabetes mellitus. However, average HbA1c was reduced from 13.3% at baseline to 9.1% at 6 months.

Unsurprisingly, BIGPIC’s community model increased linkage to care, performing better than conventional care. Although there was a 70% engagement in care at 9 months in this model, comparison data was not reported for conventional care. However, the high linkage to care with the model is similar to other studies^41^. Adopting community care approaches has resulted in better care engagement in various diseases^42^. Our finding thus suggests that adopting community models of care has the potential to improve linkage and engagement in care for diabetes management in SSA. The success seen in linkage to care and engagement in care reported within BIGPIC’s community model possibly resulted from their peer-based approach, in line with reports from other studies.^43,44^ Further, the micro-finance component which made funds available and promoted income-generating activities likely contributed to increased linkage and engagement in care.

The HDM intervention and Mobile Diab system’s eHealth model reported high acceptability of both models to both patients and health workers. Although there was no comparison of ‘acceptability’ to conventional care, several other studies report high acceptability of eHealth by end-users^45,46^. A recent study evaluating the implementation of home delivery of medication for various illness during the COVID-19 pandemic found that patients and providers preferred the continuance of this approach, with overall improvements in patient adherence to medication^47^. The eHealth model observed a clinically relevant reduction of HbA1c^48^. In a systematic review conducted to determine the effectiveness of telemedicine in the delivery of diabetes care in low- and middle-income countries^10^, telemedicine yielded significant reductions in HbA1c, with interventions via telephone and short message service yielding the highest treatment effects^10^.

Our scoping review has several limitations. First, as we did not target grey literature, we may have missed some models of care. Second, design, patient recruitment and outcome definition and evaluation differed across included studies making comparison of outcomes impossible. For instance, acceptability was assessed differently between the two studies that reported the outcome. Third, the overall number of T2DM patients enrolled in the four community-based models of care was relatively low. Fourth, lack of comparison groups in some of the included studies made it difficult to fully interpret the findings in the studies. Finally, all studies had substantial flaws in design and/or reporting making conclusions on effectiveness of community-based care for T2DM in SSA impossible.

## Conclusion

Although community-based care for patients with T2DM in SSA may be a promising approach to improve access to diagnosis and care, current evidence on such models is very limited. We identified only four studies reporting on models of care in SSA where a substantial part of the management was moved from the facility to the community. In total these four studies report on 383 patients with T2DM enrolled in one of these care models. The studies hint at opportunities and challenges community-based T2DM care may provide. However, larger scale studies, ideally randomized, with mid-to long-term outcomes on key-indicators such as engagement in care, HbA1c, and occurrence of diabetes complications are needed before a roll-out of community-based care for T2DM patients in SSA can be recommended.

## Data Availability

All relevant data are within the manuscript and its Supporting Information files.

## Declarations

### Ethics approval and consent to participate

Not applicable

### Consent for publication

Not applicable

### Availability of data and materials

All data generated and analyzed during this study are included in this article and attached Additional Files. All articles used for the review are in the references section.

Reporting guidelines: PRISMA-ScR checklist for ‘Community-based models of care for management of type 2 diabetes mellitus among non-pregnant adults in sub-Saharan Africa: a systematic scoping review,’ is available as Supplementary file, S7 table.

### Authors’ contributions

EF, LG and NDL conceptualized the study and design. JH led the literature search and de-duplication of sources. EF, LG, FU and ER conducted title/abstract and full text screening, and extracted the data. EF, FU, LG and NDL drafted the manuscript. ER, AA, FR, JMB, RG, GFD, BL, and MW reviewed the manuscript draft. All authors reviewed the results and approved the final version of the manuscript.

## Acknowledgements

We thank Christian Appenzeller from the University of Basel Library for the support in developing as well as peer reviewing the search strategy.

## Supplementary file

S1 table: Search strategy

S2 table: Inclusion and exclusion criteria

S3 table: Components of community-based care

S4 table: Data extraction tool

S5 table: Risk of bias assessment for cohort studies

S6 table: Risk of bias assessment for RCTs

S1 appendix: PRISMA-ScR checklist.

