## Supplementary file for "Community-based models of care for management of type 2 diabetes mellitus among non-pregnant adults in sub-Saharan Africa: a scoping review"

**S1 table. Search in Embase (Elsevier) conducted on 08/11/2022**

| N | Search term | | Hits |
| --- | --- | --- | --- |
| #1 | ('decentralization'/exp OR 'telehealth '/exp OR 'telemetry'/exp OR 'teleophthalmology'/exp OR Telehealth:ab,ti OR Tele-health:ab,ti OR Telemedic*:ab,ti OR Tele-medic*:ab,ti OR telemetry:ab,ti OR tele-metry:ab,ti OR biotelemetry:ab,ti OR radiotelemetry:ab,ti OR teleradiometry:ab,ti OR telesensing:ab,ti OR tele-sensing:ab,ti OR telemonitor*:ab,ti OR tele-monitor*:ab,ti OR telenurs*:ab,ti OR tele-nurs*:ab,ti OR teleconsultat*:ab,ti OR tele-consultat*:ab,ti OR telerehabilitation*:ab,ti OR tele-rehabilitation*:ab,ti OR tele-ophthalmology:ab,ti OR teleophthalmology:ab,ti OR ((virtual OR remote OR distance OR distant OR e) NEXT/2 (medic* OR monitor* OR nurs* OR consultat* OR rehabilitation* OR support* OR counsel* OR treatment* OR therap* OR sensing)):ab,ti OR (emedic* OR emonitor* OR enurs* OR econsultat* OR erehabilitation OR esupport* OR ecounsel* OR etreatment* OR etherap* OR esensing):ab,ti OR Mhealth:ab,ti OR M-health:ab,ti OR 'mobile health':ab,ti OR Ehealth:ab,ti OR E-health:ab,ti OR 'e-counseling'/exp OR 'e-mail'/exp OR 'hotline'/exp OR 'interactive voice response system'/exp OR 'internet'/de OR 'web-based intervention'/exp OR 'mobile phone'/exp OR 'telephone'/exp OR 'social media'/exp OR 'telecommunication'/exp OR 'text messaging'/exp OR 'videoconferencing'/exp OR 'web conferencing'/exp OR 'webcast'/exp OR 'podcast'/exp OR 'podcasting'/exp OR 'webinar'/exp OR 'mobile application'/exp OR 'website'/exp OR 'mobile technology'/exp OR 'e mail*':ab,ti OR 'email*':ab,ti OR (electronic NEXT/2 (mail* OR messaging)):ab,ti OR hotline*:ab,ti OR 'interactive voice response system*':ab,ti OR ((web OR online OR on-line OR internet OR video OR virtual OR tele) NEAR/3 (intervention* OR conferenc* OR communication OR seminar* OR information*)):ab,ti OR internet:ab,ti OR 'world wide web':ab,ti OR www:ab,ti OR ((mobile OR cell OR smart OR portable) NEXT/2 (phone* OR telephone*)):ab,ti OR 'social media*':ab,ti OR facebook:ab,ti OR twitter:ab,ti OR telecommunication*:ab,ti OR broadcasting:ab,ti OR 'text messag*':ab,ti OR texting:ab,ti OR teleconferenc*:ab,ti OR videoconferenc*:ab,ti OR webconferenc*:ab,ti OR webcast*:ab,ti OR podcast*:ab,ti OR webinar*:ab,ti OR SMS:ab,ti OR telephone*:ab,ti OR smartphone*:ab,ti OR ((mobile OR portable OR tablet) NEXT/2 (app OR apps OR application*)):ab,ti OR website:ab,ti OR homepage:ab,ti OR 'mobile technology':ab,ti OR 'ambulatory care'/exp OR decentrali*:ab,ti OR ((ambulatory OR dispensary OR extramural OR extra-mural OR outpatient OR 'outpatient health*' OR out-patient OR 'out-patient health*' OR out-of-office) NEXT/2 (care OR healthcare OR service* OR setting* OR monitoring* OR treatment* OR therap*)):ab,ti OR 'community care'/de OR 'community based rehabilitation'/exp OR 'community health nursing'/exp OR 'community integration'/exp OR 'community program'/exp OR 'community medicine'/exp OR 'community support'/exp OR 'community based distribution'/exp OR 'task shifting'/exp OR 'task sharing'/exp OR ((community OR 'community health*' OR district OR 'public health' OR nurse-led OR pharmacist-led OR nurse-based OR pharmacist-based OR 'social service' OR collaborative) NEXT/3 (care OR healthcare OR nurs* OR integration OR service* OR program* OR medicine OR support* OR engagement OR intervention* OR deliver* OR outreach OR distribution OR network* OR 'participatory method*' OR rehabilitation* OR resource* OR strateg* OR project* OR approach* OR testing OR screening*)):ab,ti OR 'task shar*':ab,ti OR 'task shift*':ab,ti OR 'home care'/exp OR 'home monitoring'/exp OR 'residential care'/exp OR 'adult day care'/exp OR ((adult OR home OR domicil* OR domestic OR visiting OR residential OR residence) NEAR/3 (care OR healthcare OR nurs* OR help OR assistance OR service* OR treatment* OR therap* OR 'medication review*' OR pharmac* OR monitor* OR visit* OR agenc* OR testing OR screening* OR diagnos*)):ab,ti OR homecare:ab,ti OR 'house call*':ab,ti OR 'home diagnostic test'/exp OR 'housebound patient*':ab,ti OR 'adult day care':ab,ti OR 'family service'/exp OR 'family service*':ab,ti OR 'family involve*':ab,ti OR 'family based':ab,ti OR 'family-based':ab,ti OR 'family health*':ab,ti OR 'outpatient care'/exp OR 'out-of-facilit*':ab,ti OR 'health center'/exp OR ((health OR 'health service' OR sanitary) NEXT/2 (center* OR centre* OR clinic* OR institute* OR unit* OR resort* OR facility OR facilities)):ab,ti OR 'pharmacy (shop)'/de OR 'mail order pharmacy'/exp OR 'online pharmacy'/exp OR 'apothecary':ab,ti OR 'chemist shop*':ab,ti OR 'chemist`s shop*':ab,ti OR 'pharmacies':ab,ti OR 'pharmacy':ab,ti OR 'pharmaceutical service*':ab,ti OR 'rural health care'/exp OR 'school health service'/de OR 'school health nursing'/exp OR ((rural OR school* OR workplace OR worksite OR work-place OR work-site) NEAR/3 (health* OR care OR medicine OR nurse OR nurses OR nursing OR intervention* OR program* OR approach* OR service* OR strateg*)):ab,ti OR 'self care'/exp OR 'self monitoring'/exp OR (self NEXT/2 (care* OR help OR management OR monitoring OR treatment* OR medication OR nurturance OR testing OR measur*)):ab,ti OR 'selfcare':ab,ti OR 'selfhelp':ab,ti OR 'selfmanagement':ab,ti OR 'selfmonitor*':ab,ti OR 'selftreatment':ab,ti OR 'selfmedication*':ab,ti OR 'social network'/exp OR 'social care'/exp OR ((social OR spiritual) NEXT/2 (care OR support OR network* OR service* OR work)):ab,ti OR 'support group'/exp OR 'support group*':ab,ti OR 'support program*':ab,ti OR paramedical:ab,ti OR para-medical:ab,ti OR non-health:ab,ti OR non-healthcare:ab,ti OR non-medical:ab,ti OR ((health OR healthcare) NEXT/2 ('support worker*' OR assistant* OR aid*)):ab,ti OR ((community OR district OR 'public health' OR assistan* OR aid* OR staff* OR manpower OR personnel) NEAR/4 (nurse OR nurses OR nursing OR matron*)):ab,ti OR 'midlevel health professional*':ab,ti OR 'midlevel health provider*':ab,ti OR 'paramedical personnel'/de OR 'pharmacy technician'/exp OR 'health practitioner'/exp OR 'health visitor'/exp OR 'nurse'/exp OR 'nursing assistant'/exp OR 'nursing staff'/exp OR 'paramedical profession'/exp OR 'pharmacist'/de OR 'community pharmacist'/exp OR 'health auxiliary'/exp OR ((community OR 'community based' OR village OR lay OR 'allied health' OR 'auxiliary health' OR 'health auxiliary') NEXT/3 (worker* OR provider* OR personnel OR profession* OR staff OR aide* OR supporter* OR volunteer* OR advocate* OR team* OR group* OR pharmacist*)):ab,ti OR 'barefoot doctor*':ab,ti OR 'local supervisor*':ab,ti OR 'caregiver'/exp OR 'caregiver*':ab,ti OR 'care-giver*':ab,ti OR 'peer group'/de OR 'peer education'/de OR (peer NEXT/2 (group* OR support OR relation* OR education)):ab,ti OR 'participatory research'/exp OR 'patient participation'/exp OR 'patient empowerment'/exp OR 'patient engagement'/exp OR 'patient activation'/exp OR 'participatory research':ab,ti OR 'patient involvement':ab,ti OR 'patient participation':ab,ti OR 'patient empowerment':ab,ti OR 'patient activation':ab,ti OR 'patient engagement':ab,ti OR (('community'/exp OR 'religion'/exp OR 'church*':ab,ti OR workplace/exp OR 'home'/exp OR 'school'/exp OR 'school teacher'/exp) AND ('patient care'/exp OR 'health care'/exp OR 'disease management'/de OR 'drug therapy'/exp))) | 2,572,654 | |
| #2 | ('diabetes mellitus'/exp OR 'insulin resistant diabetes mellitus'/exp OR 'diabetic patient'/exp OR 'diabetic complication'/exp OR 'diabetes education'/exp OR 'diabetes educator'/exp OR 'blood glucose monitoring'/exp OR 'glycemic control'/exp OR 'glycemic index'/exp OR 'glycemic load'/exp OR 'hyperglycemia'/exp OR 'dysglycemia'/exp OR diabet*:ab,ti OR antidiabet*:ab,ti OR 'dm 2':ab,ti OR 'dm2':ab,ti OR 'dm type 2':ab,ti OR 'dm type2':ab,ti OR niddm:ab,ti OR t2dm:ab,ti OR 't2 dm':ab,ti OR ((glycemic OR glycaemic OR glycemia OR glycaemia OR glycemie) NEXT/2 (control OR index OR value OR load)):ab,ti OR hyperglycemi*:ab,ti OR hyperglycaemi*:ab,ti OR hyperglucemi*:ab,ti OR hyper-glycemi*:ab,ti OR hyper-glycaemi*:ab,ti OR hyper-glucemi*:ab,ti OR dysglycemi*:ab,ti OR ((high OR elevated OR monitor*) NEAR/3 ('blood glucose' OR 'blood sugar' OR 'plasma glucose' OR 'plasma sugar' OR 'serum glucose' OR 'serum sugar')):ab,ti OR 'elevated blood pressure'/exp OR 'blood pressure'/de OR 'arterial pressure'/exp OR 'systolic blood pressure'/de OR 'abnormal blood pressure'/de OR 'blood pressure measurement'/exp OR 'blood pressure monitor'/exp OR 'blood pressure monitoring'/exp OR 'hypertension complications'/exp OR 'hypertensive patient'/exp OR 'antihypertensive therapy'/exp OR (hypertens* OR 'hyper tens*' OR 'blood pressur*' OR 'arterial pressur*' OR 'vascular pressur*' OR 'intravascular pressur*' OR 'blood tension' OR 'arterial tension' OR 'vascular tension' OR 'intravascular tension' OR antihypertensive):ab,ti OR 'insulin resistance'/exp OR 'insulin resistance index'/exp OR 'metabolic syndrome*':ab,ti OR 'insulin resistance':ab,ti) | 2,736,144 | |
| #3 | ('africa south of the sahara'/exp OR 'africa south of the sahara':ab,ti OR 'sub-sahara* africa*':ab,ti OR 'subsahara* africa*':ab,ti OR 'black africa*':ab,ti OR 'west africa'/exp OR 'west african'/exp OR 'west* africa*':ab,ti OR 'east african'/exp OR 'east* africa*':ab,ti OR 'southern african'/exp OR 'south* africa*':ab,ti OR 'central african'/exp OR 'central africa*':ab,ti OR 'equatorial africa*':ab,ti OR 'middle africa'/exp OR 'middle africa*':ab,ti OR angola*:ab,ti OR benin*:ab,ti OR dahomey:ab,ti OR botswana*:ab,ti OR bechuanaland:ab,ti OR 'burkina faso':ab,ti OR burkin*:ab,ti OR 'upper volta':ab,ti OR burundi*:ab,ti OR urundi:ab,ti OR cameroon*:ab,ti OR camerun:ab,ti OR kamerun:ab,ti OR cameroun:ab,ti OR 'cape verde*':ab,ti OR 'cabo verde':ab,ti OR centrafri*:ab,ti OR 'ubangi-shari':ab,ti OR 'oubangi-shari':ab,ti OR chad*:ab,ti OR tchad:ab,ti OR comoro*:ab,ti OR comores:ab,ti OR comoran:ab,ti OR comorian:ab,ti OR congo*:ab,ti OR kongo:ab,ti OR zaire:ab,ti OR 'cote d`ivoire':ab,ti OR 'ivory coast':ab,ti OR ivorian*:ab,ti OR djibouti*:ab,ti OR 'afars and issas':ab,ti OR eritrea*:ab,ti OR eswatini:ab,ti OR swazi*:ab,ti OR ethiopia*:ab,ti OR abyssinia:ab,ti OR gabon*:ab,ti OR gabun:ab,ti OR gambia*:ab,ti OR senegambia:ab,ti OR ghana*:ab,ti OR 'gold coast':ab,ti OR guinea*:ab,ti OR guinée:ab,ti OR guiné:ab,ti OR 'bissau-guinean':ab,ti OR equatoguinean:ab,ti OR kenya*:ab,ti OR lesotho*:ab,ti OR basutoland:ab,ti OR liberia*:ab,ti OR madagasca*:ab,ti OR malagasy*:ab,ti OR malawi*:ab,ti OR nyasaland:ab,ti OR mali:ab,ti OR malian*:ab,ti OR mauritania*:ab,ti OR mauritanie:ab,ti OR mauritius:ab,ti OR 'république de maurice':ab,ti OR mayotte:ab,ti OR mahoran*:ab,ti OR mozambi*:ab,ti OR mocambique:ab,ti OR namibia*:ab,ti OR niger*:ab,ti OR réunion:ab,ti OR réunionese:ab,ti OR rwanda*:ab,ti OR ruanda*:ab,ti OR senegal*:ab,ti OR 'seychellene'/exp OR seychell*:ab,ti OR 'sierra leon*':ab,ti OR somali*:ab,ti OR 'sudanese'/exp OR sudan*:ab,ti OR tanzania*:ab,ti OR tansania*:ab,ti OR tanganyika:ab,ti OR zanzibar:ab,ti OR togo*:ab,ti OR uganda*:ab,ti OR zambia*:ab,ti OR sambia*:ab,ti OR zimbabwe*:ab,ti OR rhodesia:ab,ti OR luanda:ab,ti OR lubango:ab,ti OR cabinda:ab,ti OR 'porto-novo':ab,ti OR cotonou:ab,ti OR 'abomey-calavi':ab,ti OR gaborone:ab,ti OR ouagadougou:ab,ti OR 'bobo-dioulasso':ab,ti OR bujumbura:ab,ti OR yaounde:ab,ti OR douala:ab,ti OR praia:ab,ti OR bangui:ab,ti OR n`djamena:ab,ti OR ndjamena:ab,ti OR moroni:ab,ti OR brazzaville:ab,ti OR 'pointe-noire':ab,ti OR kinshasa:ab,ti OR 'mbuji-mayi':ab,ti OR lubumbashi:ab,ti OR kananga:ab,ti OR kisangani:ab,ti OR bukavu:ab,ti OR yamoussoukro:ab,ti OR abidjan:ab,ti OR malabo:ab,ti OR asmara:ab,ti OR mbabane:ab,ti OR 'addis ababa':ab,ti OR libreville:ab,ti OR banjul:ab,ti OR accra:ab,ti OR kumasi:ab,ti OR 'sekondi takoradi':ab,ti OR conakry:ab,ti OR bissau:ab,ti OR nairobi:ab,ti OR mombasa:ab,ti OR mombassa:ab,ti OR maseru:ab,ti OR monrovia:ab,ti OR antananarivo:ab,ti OR lilongwe:ab,ti OR 'blantyre-limbe':ab,ti OR bamako:ab,ti OR nouakchott:ab,ti OR 'port louis':ab,ti OR maputo:ab,ti OR matola:ab,ti OR nampula:ab,ti OR windhoek:ab,ti OR niamey:ab,ti OR abuja:ab,ti OR lagos:ab,ti OR kano:ab,ti OR ibadan:ab,ti OR 'port harcourt':ab,ti OR kigali:ab,ti OR 'sao tome*':ab,ti OR dakar:ab,ti OR freetown:ab,ti OR mogadishu:ab,ti OR hargeisa:ab,ti OR hargeysa:ab,ti OR pretoria:ab,ti OR 'cape town':ab,ti OR johannesburg:ab,ti OR Soweto:ab,ti OR durban:ab,ti OR 'port elizabeth':ab,ti OR 'west rand':ab,ti OR juba:ab,ti OR khartoum:ab,ti OR nyala:ab,ti OR 'dar es salaam':ab,ti OR dodoma:ab,ti OR mwanza:ab,ti OR lome:ab,ti OR kampala:ab,ti OR lusaka:ab,ti OR harare:ab,ti) | 584,564 | |
| #4 | #1 AND #2 AND #3 | 4,983 | |
| #5 | (('animal'/de OR 'animal experiment'/exp OR 'nonhuman'/de) NOT ('human'/exp OR 'human experiment'/de)) | 6,993,505 | |
| #6 | ('juvenile'/exp NOT 'adult'/exp) | 2,716,368 | |
| #7 | [conference abstract]/lim | 4,589,213 | |
| #8 | #5 OR #6 OR #7 | 13,496,979 | |
| #9 | **#4 NOT #8** | **3,701** | |

S2 table. Inclusion and exclusion criteria

| Parameter | Inclusion criteria | Exclusion criteria |
| --- | --- | --- |
| Population | - Individuals aged 18 years and above, all genders, ethnic groups, education levels, socio-economic levels - Diagnosed with T2DM using the standard diagnostic criteria - In any of Angola, Benin, Botswana, Burkina Faso, Burundi, Cameroon, Central African Republic, Chad, Congo, Cote d'Ivoire, Equatorial New Guinea, Eritrea, Ethiopia, eSwatini, Gabon, Gambia, Ghana, Guinea, Guinea-Bissau, Kenya, Lesotho, Liberia, Madagascar, Malawi, Mali, Mauritania, Mauritius, Mozambique, Namibia, Niger, Nigeria, Rwanda, Senegal, Sierra Leone, Somalia, South Africa, Sudan (North, South), United Republic of Tanzania, Togo, Uganda, Zaire, Zambia, Zimbabwe. | Individuals diagnosed as having impaired glucose tolerance, pregnant women |
| Intervention | Community-based care, that is patient care different from the traditional facility-based model considering the location, frequency of contact with care provider and cadre of staff (see S3 Table) |  |
| Comparator | Traditional facility-based care, where available. |  |
| Outcomes | Studies reporting at least one the following outcomes will be included:   - Clinical outcomes: of interest are tasting blood glucose, random blood glucose, glycated haemoglobin (HbA1c), episodes of hypoglycaemia and hyperglycaemia, adherence to T2DM medication, development of complications like retinopathy, nephropathy, diabetic foot syndrome, cardiovascular diseases and cerebrovascular diseases - Engagement in care - Acceptability to patients or providers | Studies not reporting any of the outcomes |
| Study design | - Prospective or retrospective cohorts - Randomised control trials - Non-randomised control trials - Quasi-randomised control trials - Systematic or other reviews (to screen for additional original articles) | Treatment guidelines, mathematical models, editorials, viewpoints, commentaries |
| Timing | None |  |
| Sector | Services to the general public provided and or managed by government health infrastructure, or through non-governmental organisations |  |
| Required descriptive data about model | - Population/target groups - Type of patients - Community site - Health provider cadre - Frequency of service - Other services provided within the same care-model, e.g arterial hypertension, HIV, tuberculosis | - Incomplete information that impedes full model characterization and definition |

S3 table. components of community-based model of care

| **WHO** | - Any professional and non-professional cadre - Doctors, medical non-physician clinicians ,nurses, pharmacists, community health workers (and similar), peers, self-care, psychologists and social workers, family members - Traditional healers (community members not providing western ,health care - If non-professional providers: whether the project provides (or not) supervision and training from medical providers (inclusion criteria). |
| --- | --- |
| **POPULATION** | - Individuals who screen positive for T2DM. |
| **WHERE** | - Outside of the compound of a permanent health care facility. This may include, but not restricted to: community-based settings: outreach services, home-based care, places used for gathering (religious centres, schools, markets, shops) or delivering other services to citizens. Also, it includes e-health interventions. |
| **HOW OFTEN** | - Model foresees a reduction in number of patients visits to the permanent health facility, as compared to the standard of care. - The community part should not be an add-on to the care at the facility, but substitute some of the patient’s contact with facilities. |
| **WHAT** | Treatment provision in the community should include one of the following components:   - Long-term medication prescription/distribution - Point of care monitoring (eg. with glucometer) - Long-term lifestyle change support (at least 1 follow up encounter with a care provider)   The following elements may be part of the model and will be described:   - Diagnosis of chronic complications - Pharmaceutic treatment - Screening and early diagnosis of disease - Rehabilitation - Behavioral interventions, health promotion, education |

S4 table. Data extraction tool.

| **Community-based models of care for management of type 2 diabetes mellitus among non-pregnant adults in sub-Saharan Africa**  **Data extraction form** | |
| --- | --- |
| I. Study ID information | |
| Study ID | First Name/Publication Year |
| First Author | Name |
| Was author contacted? | 1 – Yes 2 – No If yes, dates(s) |
| If yes, author response? |  |
| Study data | 1 - Published 2 - In-press 3 – Ongoing  4 – Not published (gray literature) |
| Title |  |
| Year (of publication) | YYYY |
| Year study start date | YYYY or 9 – Not reported |
| Language | 1 – English 2 – Other If other, specify: |
| II. Study details | |
| Country (ies) where study was conducted  List all |  |
| Study design | 1 – Randomized controlled trial 2 – Cross-sectional 3 – Cohort 4 – Mixed-methods  5- Other, specify 9 – Could not tell  If other, describe |
| Participant selection | 1 - Consecutive  2 - Random  3 - Convenience  4 - Other  9 - Unknown/Not reported |
| Direction of study data collection | 1 – Prospective 2 – Retrospective ( period data was collected in months)  3- Cross-sectional 9 – Unknown/Not reported |
| Dates of data collection | 1-From MM/YY to MM/YY  2- Other, specify 9 – Could not tell |
| Duration of follow up (months) |  |
| Inclusion criteria in study (related to which outcome authors report, and community care described) | 1. Blood sugar control 2. Retention in care 3. End-organ damage 4. Acceptability 5. Other 9. Unknown/Not reported 10. Community care described: |
| III. Participants and setting characteristics | |
| ***If study has no intervention or control groups, fill ‘control’ column*** \| Intervention \| Control; Age(mean, SD) \| \|; Sex, female % \| \| Not described Others: | |
| Community setting of study (as defined by authors) | 0 – Urban 1 – Semiurban 2 – Rural 9 – Unknown/Not reported |
| Number of participants recruited (add) | Total:  Comments: |
| Participants recruitment technique | 1. Local drug shops 2. Community health services 3. Community pharmacies 4. Households 5. Others. Describe:   9 – Unknown/Not reported |
| Participants inclusion/ exclusion criteria | 6. |
| Length of follow up (months) |  |
| Ethnicity? |  |
| VI. Intervention | |
| Location of service delivery | 0 – Household 1 – Pharmacy  2 – Outreach services  3- Places used for gathering (religious centers, schools, markets, shops)  4- eHealth (distance support)  9 – Unknown/Not reported |
| Frequency of interaction (described) | 1 – Yes. Describe:  9 – Unknown/Not reported |
| Health care worker providing service (professional cadre) | 1. Doctor 2. Medical NPCs (non-physician clinicians) 3. Nurse 4. Pharmacists (or similar) 5. Community health workers (and similar) 6. Peers 7. Self-care 8. Family members 9. Traditional healers (community members not providing western (?) health care   0-Unknown/Not reported |
| Preparation/training for HCWs providing services | 1 Specific training 0 Unknown/Not reported |
| MINIMUM component of care included | 1. Long-term medication prescription/distribution  2. POC monitoring (eg. with glucometer) 3. Long-term lifestyle change support (at least 1 follow up encounter with a HCW) |
| OTHER component of care included | 1. Diagnosis of chronic complications 2. Initial medication 3. Screening and early diagnosis of disease  4. Rehabilitation  5. Behavioural interventions, health promotion, education  0 Unknown/Not reported |
| Type of treatment | 1. Metformin 2. Sulfonylureas 3. Meglitinides  4. Thiazolidinediones 5. DPP-4 Inhibitors 6. Others(including combinations) 9 – Unknown/Not reported |
| Type of ehealth tool used? | 1.Remote support  2.SMS  3. Others (specify) |
| V. Outcomes | |
| ***if study has not intervention and control arms, fill only ‘control’ column*** \| Intervention \| Control; – FBS values \| \|; 2 – HbA1c \| \|; 3 – Self-reported adherence to treatment \| \|; 4. side effects to medication \| \|; 5. Feasibility (based on reaching outcomes) \| \|; 6.Engagement in care (as defined by authors) \| \|; 7.End organ damage \| \|; 8. Acceptability \| \| | |

| **Author and publication year** | **Selection** | | | | **Comparability** | | **Outcome** | | | **Score** | **Quality** |
| --- | --- | --- | --- | --- | --- | --- | --- | --- | --- | --- | --- |
|  | **1** | **2** | **3** | **4** | **1** | | **1** | **2** | **3** |  |  |
| Neal et al. 2022 | - | 🗸 | - | 🗸 | 🗸 | | - | 🗸 | 🗸 | 5 | High risk |
| Pastakia et. al. 2017 | 🗸 | - | 🗸 | 🗸 | - | - | | 🗸 | - | 4 | High risk |
| Ndou et. al 2013 | - | 🗸 | 🗸 | 🗸 | - | - | | - | 🗸 | 4 | High risk |

S5 Table. Risk of bias assessment for cohort studies (Newcastle-Ottawa Scale*)

**Items marked with a tick (🗸) for each category are counted as one (“1”) and factored into the final scoring which ranges from zero to nine (lowest to highest). To simplify interpretation, studies that scored less than 6 are categorized as being of high risk of bias.*

| **Author and publication year** | **Bias arising from the randomization process** | **Bias due to deviations from intended interventions** | **Bias due to missing outcome data** | **Bias in measurement of the outcome** | **Bias in selection of the reported result** | **Overall bias** |
| --- | --- | --- | --- | --- | --- | --- |
| Takenda et. al. 2014 | High risk | Some concerns | Low risk | Some concerns | Some concerns | High risk^*^ |

S6 Table. Risk of bias assessment for RCT (Cochrane Collaboration’s tool)

**Overall risk of bias judgement is given as high risk, if the study is judged to be of high risk in at least one domain; some concerns, if the study raised some concerns in at least one domain; and low risk of bias if the study was judged to be of low risk in all domains.*

**Preferred Reporting Items for Systematic reviews and Meta-Analyses extension for Scoping Reviews (PRISMA-ScR) Checklist**

S1 Appendix. PRISMA-ScR checklist for Community-based models of care for management of type 2 diabetes mellitus among non-pregnant adults in sub-Saharan Africa: a systematic scoping review

| **SECTION** | **ITEM** | **PRISMA-ScR CHECKLIST ITEM** | **REPORTED ON PAGE #** |
| --- | --- | --- | --- |
| **TITLE** | | | |
| Title | 1 | Identify the report as a scoping review. | 1 |
| **ABSTRACT** | | | |
| Structured summary | 2 | Provide a structured summary that includes (as applicable): background, objectives, eligibility criteria, sources of evidence, charting methods, results, and conclusions that relate to the review questions and objectives. | 2-3 |
| **INTRODUCTION** | | | |
| Rationale | 3 | Describe the rationale for the review in the context of what is already known. Explain why the review questions/objectives lend themselves to a scoping review approach. | 4-5 |
| Objectives | 4 | Provide an explicit statement of the questions and objectives being addressed with reference to their key elements (e.g., population or participants, concepts, and context) or other relevant key elements used to conceptualize the review questions and/or objectives. | 5 |
| **METHODS** | | | |
| Protocol and registration | 5 | Indicate whether a review protocol exists; state if and where it can be accessed (e.g., a Web address); and if available, provide registration information, including the registration number. | 5 |
| Eligibility criteria | 6 | Specify characteristics of the sources of evidence used as eligibility criteria (e.g., years considered, language, and publication status), and provide a rationale. | 5,6 |
| Information sources* | 7 | Describe all information sources in the search (e.g., databases with dates of coverage and contact with authors to identify additional sources), as well as the date the most recent search was executed. | 6 |
| Search | 8 | Present the full electronic search strategy for at least 1 database, including any limits used, such that it could be repeated. | 5 |
| Selection of sources of evidence† | 9 | State the process for selecting sources of evidence (i.e., screening and eligibility) included in the scoping review. | 6-7 |
| Data charting process‡ | 10 | Describe the methods of charting data from the included sources of evidence (e.g., calibrated forms or forms that have been tested by the team before their use, and whether data charting was done independently or in duplicate) and any processes for obtaining and confirming data from investigators. | 6-7 |
| Data items | 11 | List and define all variables for which data were sought and any assumptions and simplifications made. | 6 |
| Critical appraisal of individual sources of evidence§ | 12 | If done, provide a rationale for conducting a critical appraisal of included sources of evidence; describe the methods used and how this information was used in any data synthesis (if appropriate). | 7 |
| Synthesis of results | 13 | Describe the methods of handling and summarizing the data that were charted. | 7 |
| **RESULTS** | | | |
| Selection of sources of evidence | 14 | Give numbers of sources of evidence screened, assessed for eligibility, and included in the review, with reasons for exclusions at each stage, ideally using a flow diagram. | 7, 10 |
| Characteristics of sources of evidence | 15 | For each source of evidence, present characteristics for which data were charted and provide the citations. | 8 |
| Critical appraisal within sources of evidence | 16 | If done, present data on critical appraisal of included sources of evidence (see item 12). | 12 |
| Results of individual sources of evidence | 17 | For each included source of evidence, present the relevant data that were charted that relate to the review questions and objectives. | 8- 12 |
| Synthesis of results | 18 | Summarize and/or present the charting results as they relate to the review questions and objectives. | 8-12 |
| **DISCUSSION** | | | |
| Summary of evidence | 19 | Summarize the main results (including an overview of concepts, themes, and types of evidence available), link to the review questions and objectives, and consider the relevance to key groups. | 13-14 |
| Limitations | 20 | Discuss the limitations of the scoping review process. | 14 |
| Conclusions | 21 | Provide a general interpretation of the results with respect to the review questions and objectives, as well as potential implications and/or next steps. | 15 |
| **FUNDING** | | | |
| Funding | 22 | Describe sources of funding for the included sources of evidence, as well as sources of funding for the scoping review. Describe the role of the funders of the scoping review. | 15 |

JBI = Joanna Briggs Institute; PRISMA-ScR = Preferred Reporting Items for Systematic reviews and Meta-Analyses extension for Scoping Reviews.

* Where *sources of evidence* (see second footnote) are compiled from, such as bibliographic databases, social media platforms, and Web sites.

† A more inclusive/heterogeneous term used to account for the different types of evidence or data sources (e.g., quantitative and/or qualitative research, expert opinion, and policy documents) that may be eligible in a scoping review as opposed to only studies. This is not to be confused with *information sources* (see first footnote).

‡ The frameworks by Arksey and O’Malley (6) and Levac and colleagues (7) and the JBI guidance (4, 5) refer to the process of data extraction in a scoping review as data charting*.*

§ The process of systematically examining research evidence to assess its validity, results, and relevance before using it to inform a decision. This term is used for items 12 and 19 instead of "risk of bias" (which is more applicable to systematic reviews of interventions) to include and acknowledge the various sources of evidence that may be used in a scoping review (e.g., quantitative and/or qualitative research, expert opinion, and policy document).

*From:* Tricco AC, Lillie E, Zarin W, O'Brien KK, Colquhoun H, Levac D, et al. PRISMA Extension for Scoping Reviews (PRISMAScR): Checklist and Explanation. Ann Intern Med. 2018;169:467–473. [doi: 10.7326/M18-0850](http://annals.org/aim/fullarticle/2700389/prisma-extension-scoping-reviews-prisma-scr-checklist-explanation).
